# Factors Associated with Longitudinal Psychological and Physiological Stress in Health Care Workers During the COVID-19 Pandemic

**DOI:** 10.1101/2020.12.21.20248593

**Authors:** Robert P. Hirten, Matteo Danieletto, Lewis Tomalin, Katie Hyewon Choi, Micol Zweig, Eddye Golden, Sparshdeep Kaur, Drew Helmus, Anthony Biello, Renata Pyzik, Claudia Calcogna, Robert Freeman, Bruce E Sands, Dennis Charney, Erwin P Bottinger, Laurie Keefer, Mayte Suarez-Farinas, Girish N. Nadkarni, Zahi A. Fayad

**Affiliations:** The Dr. Henry D. Janowitz Division of Gastroenterology, Icahn School of Medicine at Mount Sinai, New York, NY, USA; The Hasso Plattner Institute for Digital Health at the Mount Sinai, New York, NY, USA; Department of Genetics and Genomic Sciences, Icahn School of Medicine at Mount Sinai, New York, NY, USA; Center for Biostatistics, Department of Population Health Science and Policy, Icahn School of Medicine at Mount Sinai; The BioMedical Engineering and Imaging Institute, Icahn School of Medicine at Mount Sinai, New York, NY, USA; Department of Population Health Science and Policy, Icahn School of Medicine at Mount Sinai, New York, New York, USA; Institute for Healthcare Delivery Science, Icahn School of Medicine at Mount Sinai, New York, New York, USA; Office of the Dean, Icahn School of Medicine at Mount Sinai; Nash Family Department of Neuroscience, Icahn School of Medicine at Mount Sinai; The Department of Psychiatry, Icahn School of Medicine at Mount Sinai, New York, NY, USA; The Department of Medicine, Icahn School of Medicine at Mount Sinai, New York, NY, USA; The Charles Bronfman Institute for Personalized Medicine, Icahn School of Medicine at Mount Sinai, New York, NY, USA; Department of Diagnostic, Molecular and Interventional Radiology, Icahn School of Medicine at Mount Sinai

## Abstract

**Introduction:** The Coronavirus Disease 2019 (COVID-19) pandemic has resulted in psychological distress in health care workers (HCWs). There is a need to characterize which HCWs are at increased risk of psychological sequela from the pandemic.

**Methods:** HCWs across seven hospitals in New York City were prospectively followed in an ongoing observational digital study using the custom Warrior Watch Study App. Participants wore an Apple Watch for the duration of the study measuring HRV throughout the follow up period. Surveys were obtained daily.

**Results:** Three hundred and sixty-one HCWs were enrolled. Multivariable analysis found New York City COVID-19 case count to be significantly associated with increased longitudinal stress (p=0.008). A non-significant decrease in stress (p=0.23) was observed following COVID-19 diagnosis, though there was a borderline significant increase following the 4-week period after a COVID-19 diagnosis via nasal PCR (p=0.05). Baseline emotional support, baseline quality of life and baseline resilience were associated with decreased longitudinal stress (p<0.001). Baseline resilience and emotional support were found to buffer against stressors, with a significant reduction in stress during the 4-week period after COVID-19 diagnosis observed only in participants in the highest tertial of emotional support and resilience (effect estimate −0.97, p=0.03; estimate −1.78, p=0.006). A significant trend between New York City COVID-19 case count and longitudinal stress was observed only in the high tertial emotional support group (estimate 1.22, p=0.005), and was borderline significant in the high and medium resilience tertials (estimate 1.29, p=0.098; estimate 1.14, p=0.09). Participants in the highest tertial of baseline emotional support and resilience had significantly reduced amplitude and acrophase of the circadian pattern of longitudinally collected heart rate variability.

**Conclusion:** Our findings demonstrate that low resilience, emotional support, and quality of life identify HCWs at risk of high perceived longitudinal stress secondary to the COVID-19 pandemic and have a distinct physiological stress profile. Assessment of HCWs for these features can identify and permit allocation of psychological support to these at-risk individuals as the COVID-19 pandemic and its psychological effects continue in this vulnerable population.

## INTRODUCTION

Increasing rates of SARS-CoV2 infections and hospitalizations, growing workloads, and concern regarding personal protective equipment have resulted in a large psychological burden on health care workers (HCWs).^1^ While prior pandemics have had psychological effects on HCWs, increasing post-traumatic stress, depression and anxiety, the scale and duration of the Coronavirus Disease 2019 (COVID-19) pandemic amplifies the risk of these adverse outcomes.^1-3^ Cross sectional studies have demonstrated that front line HCWs are at a high risk of depression, anxiety, insomnia and distress compared to the general population.^4-6^ HCWs on wards serving patients with COVID-19 reported higher levels of stress, exhaustion, depressive mood and burnout.^7, 8^ However, there is limited longitudinal data on the pandemics psychological impact on this group, limited data across health care occupations, no means to identify which HCWs are at risk of developing psychological sequela over time, and no objective evaluation of the stress response in HCWs. Identification of at risk HCWs will allow for appropriate allocation of mental health resources.

Advances in digital technology provide a means to address these limitations. Smart phone Apps can administer surveys and integrate wearable devices, such as the Apple Watch, to monitor the autonomic nervous system (ANS), a primary component of the stress response. ANS function can be ascertained through measurement of heart rate variability (HRV), a measure of the parasympathetic and sympathetic nervous systems impact on cardiac contractility through calculation of changes in the beat to beat intervals.^9^

## METHODS

### Study Design

This is an observational cohort study. The primary objective of the study was to identify characteristics associated with longitudinal stress in HCWs. The secondary aim was to determine whether changes in HRV associate with features protective against longitudinal stress development. HCWs across 7 hospitals in New York City (NYC) (The Mount Sinai Hospital, Morningside Hospital, Mount Sinai West, Mount Sinai Beth Israel, Mount Sinai Queens, New York Eye and Ear Infirmary, Mount Sinai Brooklyn) were eligible. Participants had to be current employees of one of the participating hospitals, ≥18 years of age, have an iPhone Series 6 or higher and be willing to wear an Apple Watch Series 4 or higher. An underlying autoimmune disease or the use of medications that interfere with ANS function were exclusionary.

### Study Procedures

Participants downloaded the custom Warrior Watch Study App to their iPhones and completed eligibility questions prior to signing electronic consent. Through the study App demographics, a diagnosis of anxiety or depression, perceived stress (Perceived Stress Scale-4 [PSS-4])^10^, resilience (abbreviated Connor-Davidson Resilience Scale [CD-RISC-2])^11^, emotional support (2-item PROMIS questionnaire)^12^, quality of life (2-item Global Health and Quality of Life)^13^, and optimism (Life Orientation Test)^14^ were collected at enrollment (**Supplementary Table 1**). Diagnosis of COVID-19 was defined as a positive SARS-CoV-2 nasal PCR swab reported by a study subject. Daily survey’s collect COVID-19 related symptoms and severity, degree of COVID-19 exposure at work, types of patient care at work, whether participants left their home each day, if public transportation was used, the number of people that participants interact with each day, the results of any COVID-19 nasal PCR or antibody tests, whether the subject is quarantined, if childcare needs are required and if the subject is admitted to the hospital. To enable trending of psychological well-being, subjects are prompted to complete the PSS-4 and 2-item General Health and QOL survey weekly. Participants were instructed to wear the Apple Watch for at least 8 hours per day.

### Wearable Device

The Apple Watch Series 4 or 5 was worn by subjects on the wrist to capture HRV and is connected via blue tooth to the participants iPhone. A photoplethysmogram (PPG) sensor on the Apple Watch pairs a green LED light with a light sensitive photodiode to generate time series peaks.^15^ The Apple Watch and Apple Health app calculates HRV using the standard deviation of NN intervals (SDNN) from the time differences between heart beats, categorized as the Interbeat Interval. SDNN is a time domain index reflecting sympathetic and parasympathetic nervous system activity.^9^ This is recorded by the Apple Watch during approximately 60 second recording periods (ultra-short period). All data generated by the Apple Watch are stored on the iPhone and transferred with completion of App surveys.

### Statistical Analysis

#### Survey Analyses

To account for gaps created by unanswered weekly surveys and allow comparison for each patient, we created a new chronological variable called a ‘period’. To account for participants having different time windows between each weekly survey, a period was assigned to each weekly survey according to participants’ starting and ending date. When a participant’s survey was completed less than 7 days from their previous survey date, the day after the previous survey date was regarded as the starting window date for the next period. When a participant’s survey was done 7 days or more apart from the previous survey date, the starting window date was set to 6 days prior to the current survey date. To integrate weekly psychological metrics and daily risk/health metrics, results of the daily surveys were summarized by the periods defined by the weekly surveys. Daily survey data was summarized for each period, eg: mean number of risk days per period, mean number of days left home per period, and mean symptom severity per period. To examine associations between the NYC COVID-19 case-count and perceived stress raw NYC case-count data was obtained for modelling and summarized as a mean case-count per period.^16^

#### Occupation Classification

The occupation of each participant was collected at enrollment. However, due to the pandemic, the roles, risks and responsibilities of these occupations may have changed when compared to non-pandemic job descriptions. We therefore created a new occupation metric to identify which participants were seeing patients during the study. Occupation was calculated as follows: 1) Daily clinical occupation was calculated from the daily survey where participants classified the type of patient or non-patient care responsibilities, he or she had that day. Those who reported either (a) exposure to patient areas but without patients diagnosed with COVID-19 or those being evaluated for COVID-19, or (b) exposure to areas with patients confirmed to have COVID-19 or people being investigated for COVID-19 infection, were assigned as clinical for that day. Those who responded they were at work but not caring for patients or those who were working remotely were classified as non-clinical for that day. 2) If a participant had one or more clinical days in a given period, that participant is assigned as clinical for that period. 3) If a participant has one or more clinical periods over the entire study, then they are deemed as either clinical non-trainee or clinical trainee. To be classified as a clinical trainee a participant had to be either a resident or fellow. All other occupations were classified as staff.

#### Statistical Modeling

To model longitudinal changes in stress, we used linear mixed-effect models. Fixed effects included time invariant covariates (gender, age, occupation, baseline resilience, optimism and quality of life) and time variant covariates (COVID-19 diagnosis, SARS COVID-19 antibody positive test, mobility variables). A continuous First Order Autoregressive correlation structure (over period) was found to be suitable to our data significantly increasing the likelihood function (LRT p<0.001) and leading to minimal Akaiki information criterion (AIC)/Bayesian information criterion (BIC). Model coefficients were estimated using a restricted maximum likelihood approach (REML) method using R’s *nlme* packages. Hypothesis of interest were tested using contrasts through the capabilities of the *emmeans* package.

Univariate models tested the association of each variable with longitudinal stress and identified associated factors. Variables with p<0.10 in the marginal ANOVA test were considered significant and included in the multivariate analysis. Although in univariate models random effects include only the intercept, in multivariate models, a random effect for the NYC case burden was found to be significant (LRT<0.001, lower AIC/AIC), indicating heterogeneity in the association of this variable with stress across subjects.

#### Heart Rate Variability Modelling

HRV captured from the Apple Watch demonstrated a sparse non-uniform sampling and circadian pattern making it amenable to analysis via a COSINOR model. This approach models the daily HRV circadian rhythm over a period of 24 hours which can be described using the circadian parameters: (1) Midline Statistic of Rhythm (MESOR), (2) Amplitude, and (3) Acrophase. This allows testing of the effect that model covariates have on HRV. A COSINOR model used the non-linear function Y(t) = M+*Acos*(2*π*t/*τ*+ *ϕ*) + e_i_(t), where *τ* is the period (*τ*=24h), M is the MESOR, A is the amplitude and Φ is the Acrophase. This can be transformed into the linear model *ϕ*(2*πϕ*(2*π*t/*ϕ*), with HRV written as Y(t)=M+*ϕ*x_t_ +*ϕ*z_t_ + e_i_(t). We identified a subject specific daily pattern measuring departures from this pattern as a function of emotional support, resilience, and other covariates of interest. Utilizing a mixed effect COSINOR model HRV, the introduction of random effects intrinsically models the correlation due to the longitudinal sampling. Covariates, C, were introduced as fixed effects using the equation HRV_it_ = M+*ϕ*_o_C_i_+(*ϕ*+*ϕ*_2_*ϕ*_i_*ϕ*x_it_ + (*ϕ*+*ϕ*_3_*ϕ*_i_*ϕ*z_it_ +*ϕ*+ e_i_(t). As we have described previously to test if the COSINOR curve differs between two populations of interest we performed the bootstrapping procedure where for each iteration we (1) fit a linear mixed-effect model using REWL (2) estimated the marginal means for each group defined by a covariate (3) estimated marginal means for each group using the inverse relationship, and (4) defined the bootstrapping statistics as a pairwise difference between groups.^17^ COSINOR models were used to estimate HRV MESOR, amplitude and acrophase for participants based on emotional support and resilience tertials (low, medium and high). COSINOR model covariates included time, gender, age, BMI, baseline emotional support, baseline resilience, optimism and stress, with the participant serving as a random intercept.

## RESULTS

In response to the COVID-19 pandemic we launched the Warrior Watch Study, comprised of our custom iOS App which integrates survey metrics with physiological signatures acquired from the Apple Watch. Three hundred sixty-one HCWs, characterized as any worker in a health system, were enrolled across seven hospitals in NYC in this ongoing observational study between April 29^th^ and September 29^th^, 2020, when data was censored for analysis (**Table 1**). Participants had a mean age of 37 years, were 69.3% female and were followed for a mean of 60 days (IQR 21-98 days). Clinical trainees had higher baseline resilience, compared to clinical non-trainees (p=0.03) and staff (p=0.01), higher optimism (p=0.04) and emotional support (p=0.01) compared to staff, and higher emotional support compared to clinical non-trainees (p=0.01) (**Supplementary Table 2**).

**Table 1.** Baseline demographic characteristics of the total cohort and by occupation category.

|  | **Total Cohort (n=361)** | **Staff (n=65)** | **Clinical Non-Trainee (n=217)** | **Clinical Trainee (n=40)** |
| --- | --- | --- | --- | --- |
| Age, mean (SD) | 36.8 (10.1) | 36.5 (11.0) | 37.8 (10.4) | 31.1 (3.6) |
| Body Mass Index, mean (SD) | 25.7 (5.8) |  |  |  |
| Female Gender (%) | 246 (69.3) | 43 (66.2) | 158 (73.8) | 20 (51.3) |
| Race (%) |  |  |  |  |
| Asian | 90 (24.9) | 14 (21.5) | 49 (22.6) | 14 (35.0) |
| Black | 33 (9.1) | 3 (4.6) | 23 (10.6) | 4 (10.0) |
| Other | 47 (13.0) | 7 (10.8) | 31 (14.3) | 6 (15.0) |
| White | 132 (36.6) | 26 (40.0) | 80 (36.9) | 15 (37.5) |
| Ethnicity (%) |  |  |  |  |
| Hispanic | 59 (16.3) | 15 (23.1) | 34 (15.7) | 1 (2.5) |
| Baseline Positive SARS-CoV-2 nasal PCR (%) | 22 (6.1) | 2 (3.1) | 16 (7.4) | 2 (5.0) |
| Baseline Positive SARS-CoV-2 serum antibody (%) | 35 (9.7) | 6 (9.2) | 22 (10.1) | 2 (5.0) |
| Baseline Smoking Status (%) |  |  |  |  |
| Current/Past smoker | 48 (13.5) | 10 (15.4) | 31 (14.5) | 0 (0.0) |
| Never/Rarely smoker | 307 (86.5) | 55 (84.6) | 183 (85.5) | 39 (100.0) |
| Baseline Immune Suppressing Medication (%) | 4 (1.4) | 0 (0.0) | 4 (1.9) | 0 (0.0) |
| Anxiety or Depression | 73 (20.6) | 16 (24.6) | 43 (20.1) | 7 (17.9) |
| Baseline Survey Metrics, mean (SD) |  |  |  |  |
| PSS-4 | 5.3 (3.1) | 5.5 (2.9) | 5.4 (3.1) | 5.0 (3.0) |
| CD-RISC | 5.7 (1.4) | 5.4 (1.5) | 5.7 (1.4) | 6.2 (1.3) |
| Optimism | 19.1 (4.2) | 18.4 (4.3) | 18.8 (4.2) | 20.1 (3.7) |
| Emotional Support | 6.8 (1.5) | 6.7 (1.7) | 6.8 (1.5) | 7.6 (0.9) |
| Quality of Life | 7.8 (1.5) | 7.5 (1.4) | 7.8 (1.4) | 8.0 (1.5) |
| Baseline Medical Conditions |  |  |  |  |
| Asthma | 41 (11.4) | 13 (20) | 19 (8.8) | 5 (12.5) |
| Chronic Lung Disease | 1 (0.3) | 0 (0.0) | 0 (0.0) | 1 (2.5) |
| Heart Disease | 1 (0.3) | 1 (1.5) | 0 (0.0) | 0 (0.0) |
| Cancer | 2 (0.6) | 0 (0.0) | 2 (0.9) | 0 (0.0) |
| Diabetes Mellitus | 6 (1.7) | 2 (3.1) | 4 (1.8) | 0 (0.0) |
| Hypertension | 20 (5.5) | 5 (7.7) | 11 (5.1) | 0 (0.0) |
| Pneumonia | 7 (1.9) | 1 (1.5) | 5 (2.3) | 1 (2.5) |

Univariate analysis evaluated the relationship between baseline demographics and prospectively collected survey metrics with longitudinal perceived stress (weekly PSS-4) (**Supplementary Table 3**). Baseline factors including resilience, optimism, emotional support, quality of life, male gender, and age were significantly associated with lower longitudinal stress. Baseline anxiety/depression, body mass index (BMI), weight, and asthma were significantly associated with increased longitudinal stress. Longitudinal quality of life (p<0.001) was associated with reduced longitudinal stress, while the mean number of COVID-19 cases in NYC (p=0.004) was positively associated with increased longitudinal stress. Occupation classification (staff vs clinical non-trainee, p=0.81; staff vs clinical trainee, p=0.15; clinical non-trainee vs clinical trainee, p=0.17), mean number of days caring for patients (p=0.88) and treatment of patients with COVID-19 (p=0.73) were not associated with longitudinal stress. We observed a significant reduction of stress during the 4-week period following diagnosis (p=0.014) and over the follow up period (p=0.04). Multivariable analysis found only NYC COVID-19 case count to be significantly associated with increased longitudinal stress (p=0.008). The drop in stress during the 4 week period following COVID-19 diagnosis was not significant (p=0.23), however we noted a borderline significant increase in stress following the 4-week period after a COVID-19 diagnosis (p=0.05). Baseline emotional support, baseline quality of life and baseline resilience were associated with decreased longitudinal stress (p<0.001) (**Figure 1**).

**Figure 1.**
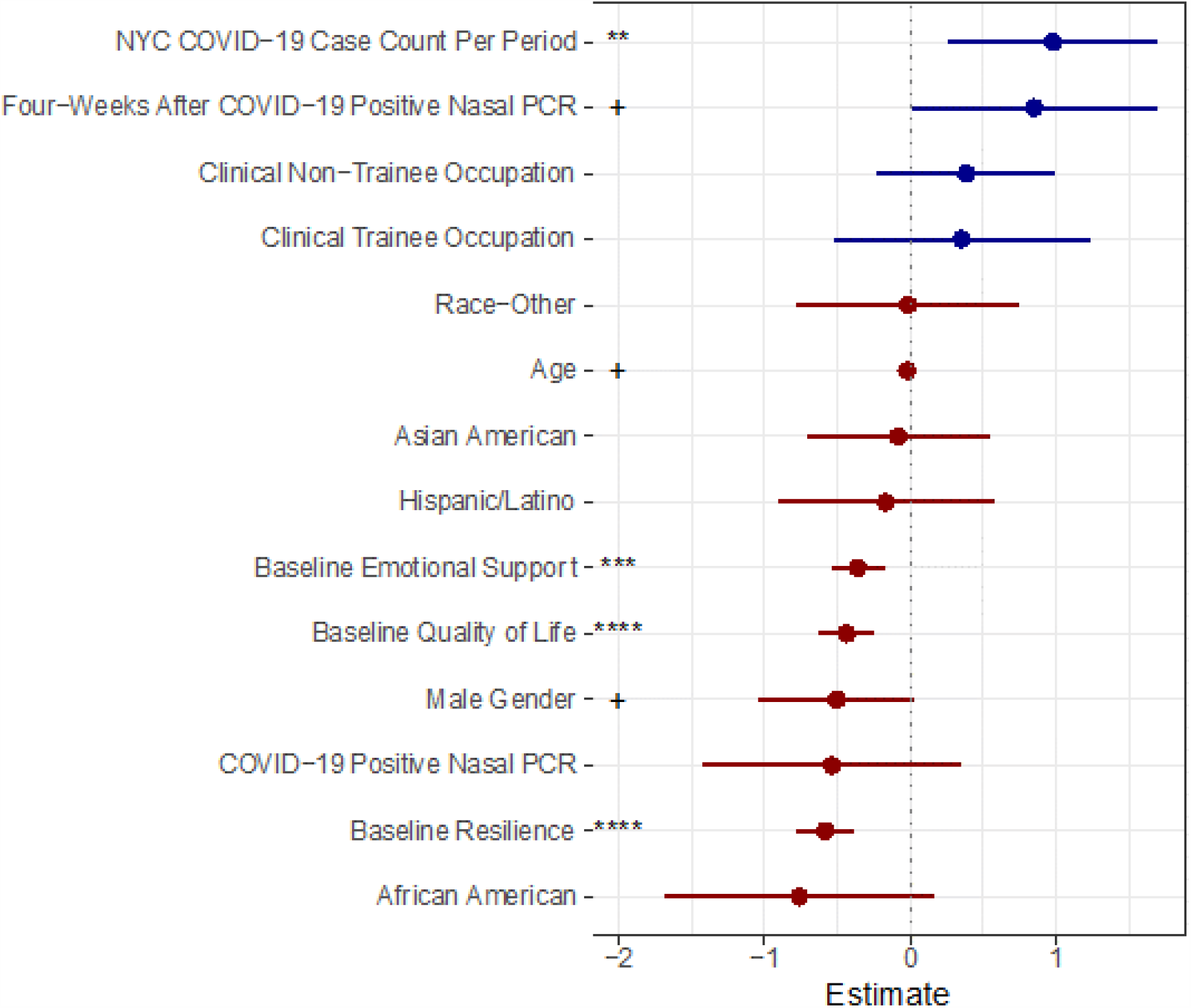
Multivariate analysis of factors associated with longitudinal stress. The scatter plot shows estimated coefficients (±confidence intervals) for variables used in the multivariate analysis. Stars indicate that variable has significant (p<0.05) association with longitudinal stress while crosses indicate a borderline significant relationship (p<0.10). Positive association is indicated in blue, negative association in red.

NYC COVID-19 case count and the 4-week period after a COVID-19 diagnosis via nasal PCR were further explored in the context of emotional support and resilience. Participants were stratified into emotional support tertials (low, medium, high). A significant reduction in stress during the 4-week period after COVID-19 diagnosis occurred only in participants in the highest tertial of emotional support (effect estimate - 0.97, p=0.03), but not in the medium (effect estimate −0.62, p=0.48), and low tertials (effect estimate 0.08, p=0.93) (**Figure 2A**). A significant trend between COVID-19 case count in NYC and longitudinal stress was observed only in the high tertial emotional support group (estimate 1.22, p=0.005), not in the low (estimate −1.45, p=0.26) or medium (estimate 0.98, p=0.16) tertials (**Figure 2b**). Stratification of the cohort into tertials for resilience demonstrated a significant reduction in stress during the 4-week period after COVID-19 diagnosis via nasal PCR in the high (estimate −1.78, p=0.006) but not medium (estimate 0.33, p=0.64) and low tertials (estimate −0.60, p=0.25) (**Figure 2c**). The impact of COVID-19 case counts in NYC demonstrated a borderline significant relationship with stress in the medium (estimate 1.29, p=0.098) and high (estimate 1.14, p=0.09), but not in the low resilience group (estimate 0.72, p=0.21) (**Figure 2d**).

**Figure 2.**
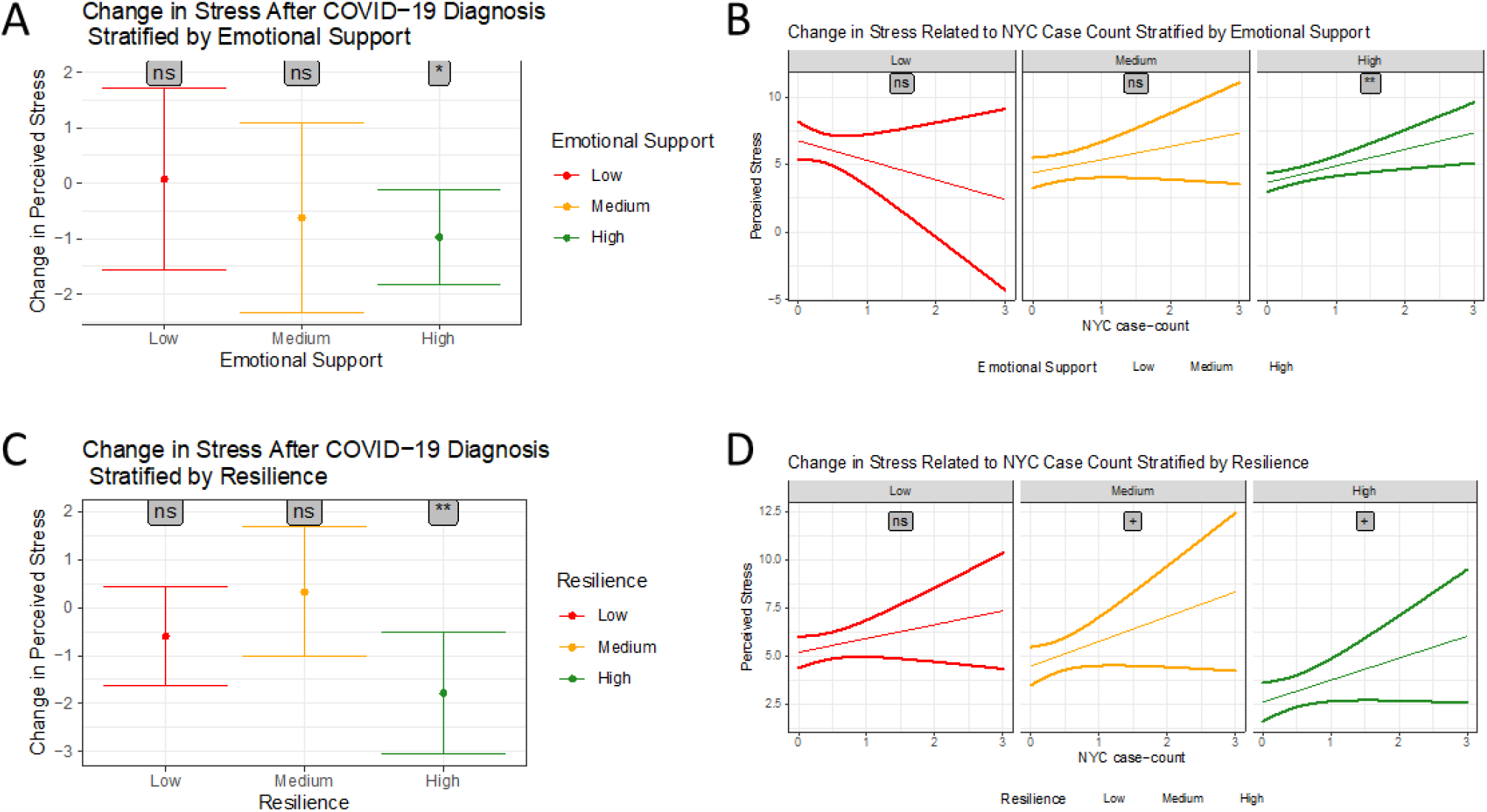
Plots (A, C) show changes in longitudinal stress following a positive COVID-19 nasal test in participants with low, medium and high emotional support (A) or resilience (C), stars indicate that change in longitudinal stress was significantly different from zero. Line plots (B, D) show relationship between New York City COVID-19 case-count and mean longitudinal stress (± Confidence Intervals) for participants with low, medium and high emotional support (B) or resilience (D), stars indicate significant trend between New York City case-count and longitudinal stress. (+p<0.1, *p<0.05, **p<0.01, ***p<0.001).

To evaluate whether the stress buffering effect of emotional support and resilience resulted in physiological differences in the stress response of HCWs we fit a COSINOR model evaluating differences in HRV (SDNN) (**Supplementary Table 4**). Significant reduction in the amplitude and acrophase of the circadian pattern of longitudinal SDNN was observed between participants with high compared to medium (p<0.001; p<0.001) and low (p=.008; p=0.004) baseline emotional support, respectively (**Figure 3a and 3b**). Significant changes in the circadian pattern of SDNN was also observed when the cohort was stratified based on baseline resilience (**Figure 3c and 3d**). The amplitude and acrophase of the circadian pattern of SDNN was significantly lower in subjects with high resilience compared to those with low (p<0.001; p=0.048) and medium (p<0.001; p<0.001) resilience, respectively (**Supplementary Table 5**).

**Figure 3.**
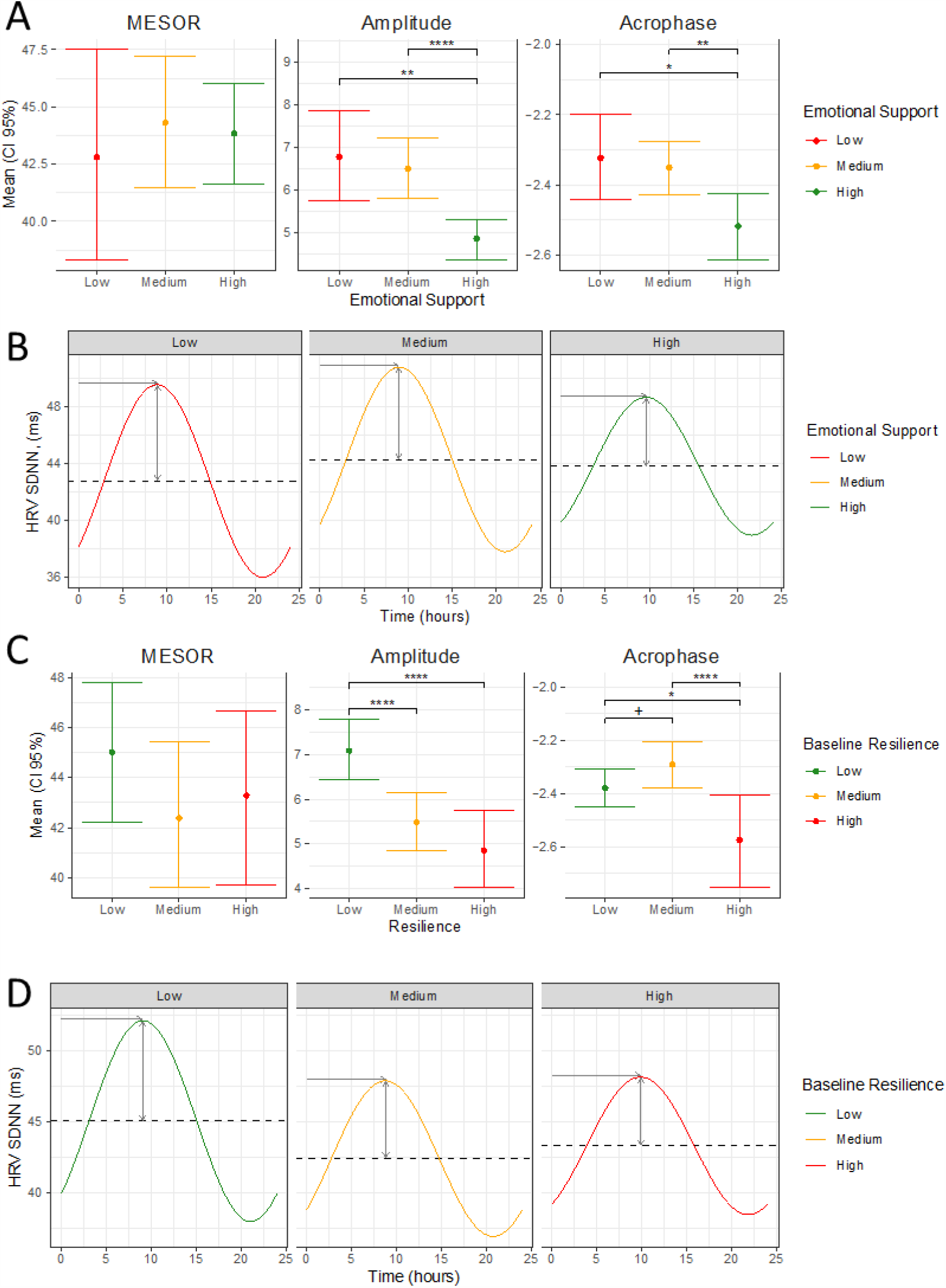
Exploring the relationship between HRV, emotional support and resilience. Plots (A, C) show mean (± 95% Confidence Intervals) HRV MESOR, Amplitude and Acrophase for participants with low, medium and high emotional support (A) or resilience (C). Stars indicate significant differences between groups. Plots (B, D) show average daily circadian HRV rhythm for participants with low, medium and high emotional support (B) or resilience (D). (+p<0.1, *p<0.05, **p<0.01, ***p<0.001).

## DISCUSSION

In summary, we conducted the first study to identify HCW characteristics that correlate with longitudinal stress during the COVID-19 pandemic and identify employees at risk of psychological sequela. We found worsening longitudinal stress is associated with the number of COVID-19 cases in the community, highlighting the effect of the environmental stressor. Baseline emotional support, resilience and quality of life defined which HCWs were prone to perceived longitudinal stress, not occupation class, and characterized a unique ANS stress profile.

In line with our findings, prior work shows that emotional support and resilience buffer against stress.^18, 19^ Resilience, defined as a reduced vulnerability to environmental stressors and the ability to overcome difficulty, is crucial to establishing social relationships and is tied to social support, which also acts as an environmental protective factor against adversity.^20-22^ In addition to demonstrating their stress protective effect in multivariate analysis, when we further evaluated NYC COVID-19 case count, a factor associated with longitudinal stress over time, we again found that those with lower emotional support or resilience were vulnerable with a dynamic stress response uncoupled from the environmental COVID-19 stressor. Similarly, the transient reduction in stress that occurs after a COVID-19 diagnosis only occurs in those with high emotional support and resilience.

A strength of our study is the objective assessment of this observation through longitudinal HRV measurements. HRV is a marker of the physiological stress response on the ANS.^23^ We found that participants with high emotional support or resilience have a physiologically distinct ANS profile demonstrating the impact of these characteristics on physiological metrics of stress. The multiple dimensions in which we reaffirmed the importance of these features substantiates their effect on longitudinal stress in HCWs. One of these features, resilience, is modifiable through targeted interventions, providing an opportunity to increase it in HCWs with low resilience. Several resilience building interventions have demonstrated to be effective in HCWs,^24, 25^ however, our findings linking HRV alterations with degree of resilience, makes HRV focused resilience building exercises an attractive option.^26^

Strengths of the study are its multicenter longitudinal study design. Furthermore, the number and type of longitudinal variables we capture allows for a robust multivariate analysis. Lastly, the incorporation of ANS parameters provide an objective assessment of the stress response. However, there are several limitations to our study. The Apple Watch provides HRV data in one-time dimension (SDNN) limiting evaluation of other metrics with outcomes of interests. The Apple Watch also provides HRV sampling sporadically throughout the day. While our modelling accounts for this, a denser sampling would allow expanded analyses.

Our findings demonstrate that low resilience, emotional support, and quality of life identify HCWs at risk of perceived and physiological longitudinal stress secondary to the COVID-19 pandemic. Assessment of HCWs for these features can permit allocation of psychological support to at-risk individuals as the COVID-19 pandemic and its psychological effects continue in this vulnerable population.

## Data Availability

This study followed health care workers. Due to the sensitive nature of their data there are limitations on the public availability of the data.

---

**Supplementary Table 1.**
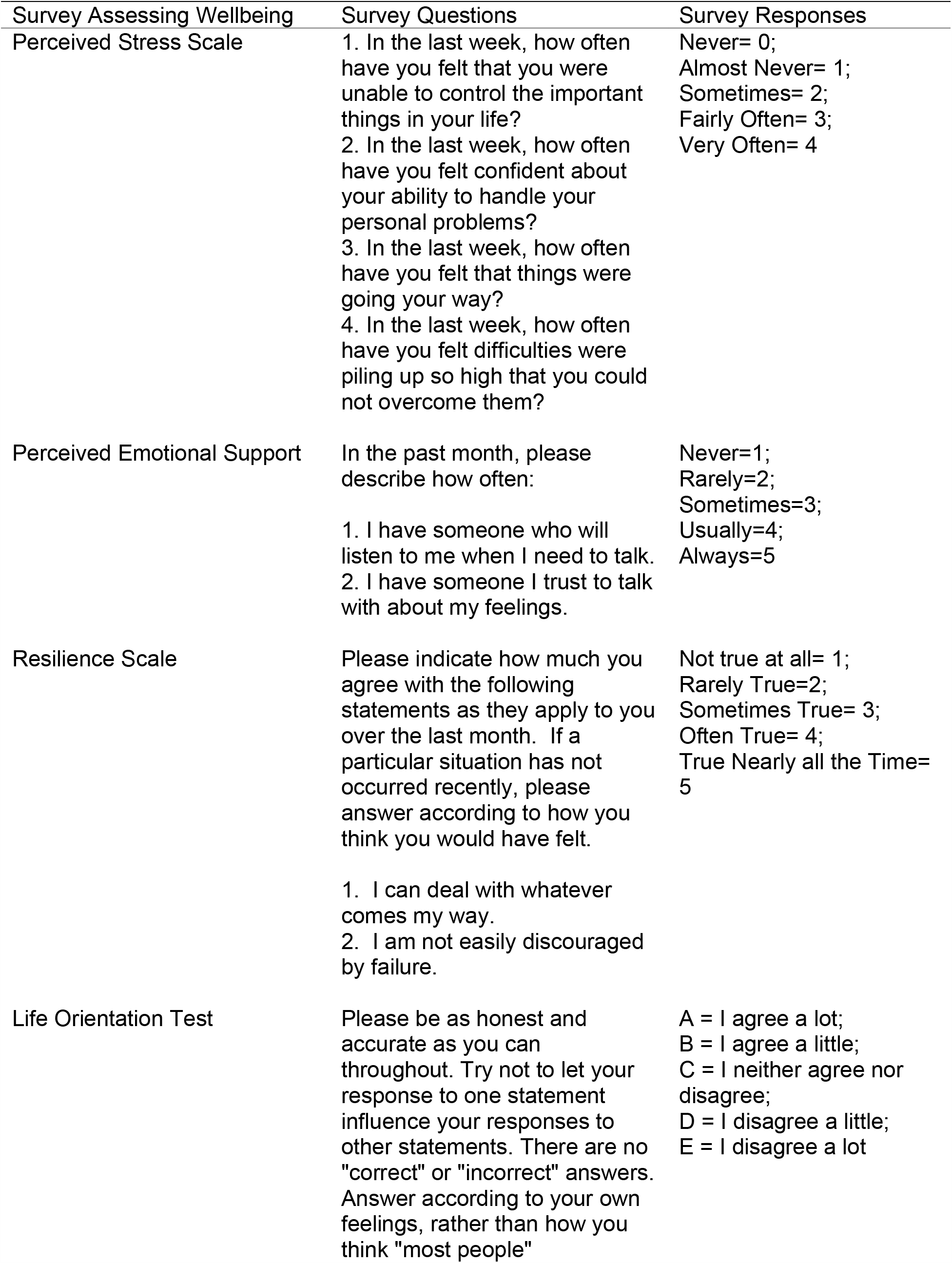
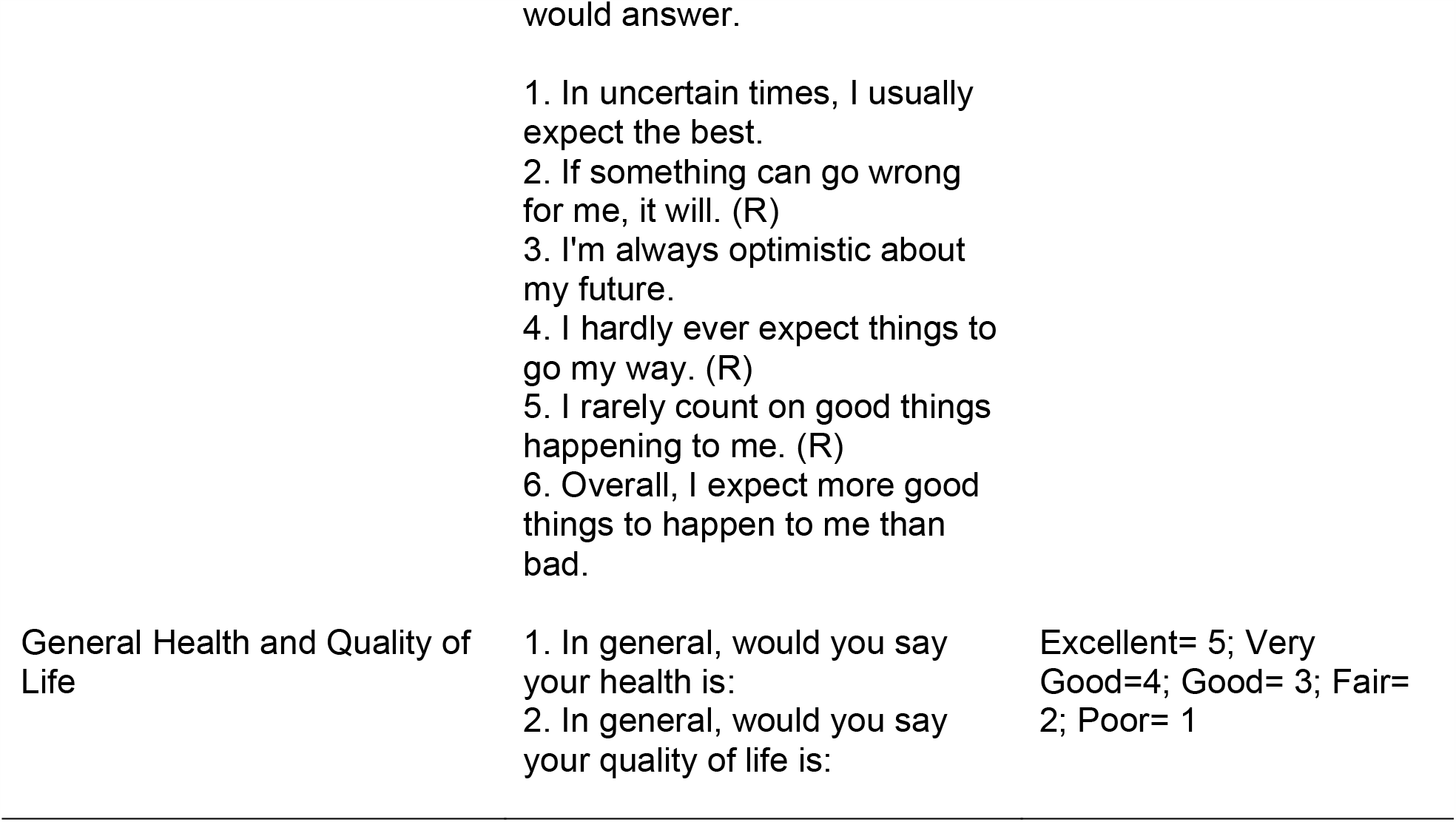
Surveys assessing psychological well-being

**Supplementary Table 2.** Statistical comparison of baseline demographic characteristics.

|  | Effect Estimate | P Value |
| --- | --- | --- |
| Age |  |  |
| Staff vs Clinical Non-Trainee | -1.343 | **0.01** |
| Clinical Non-Trainee vs Clinical Trainee | 6.673 | **<0.001** |
| Staff vs Clinical Trainee | 5.330 | 0.35 |
| Body Mass Index |  |  |
| Staff vs Clinical Non-Trainee | 0.943 | 0.25 |
| Clinical Non-Trainee vs Clinical Trainee | 3.107 | **0.01** |
| Staff vs Clinical Trainee | 4.050 | **0.01** |
| Gender Across Occupations* |  | **0.02** |
| Race Across Occupations* |  | 0.16 |
| Positive SARS-CoV-2 nasal PCR Across Occupations * |  | 0.43 |
| Positive SARS-CoV-2 serum antibody Across Occupations * |  | 0.59 |
| Smoking Status* |  | **0.04** |
| Immune Suppressing Medication* |  | 0.37 |
| Anxiety or Depression* |  | 0.66 |
| PSS-4 |  |  |
| Staff vs Clinical Non-Trainee | 0.041 | 0.93 |
| Clinical Non-Trainee vs Clinical Trainee | 0.418 | 0.44 |
| Staff vs Clinical Trainee | 0.459 | 0.47 |
| CD-RISC |  |  |
| Staff vs Clinical Non-Trainee | -0.255 | 0.20 |
| Clinical Non-Trainee vs Clinical Trainee | -0.510 | **0.03** |
| Staff vs Clinical Trainee | -0.764 | **0.01** |
| Optimism |  |  |
| Staff vs Clinical Non-Trainee | -0.439 | 0.46 |
| Clinical Non-Trainee vs Clinical Trainee | -1.28 | 0.08 |
| Staff vs Clinical Trainee | -1.72 | **0.04** |
| Emotional Support |  |  |
| Staff vs Clinical Non-Trainee | -0.131 | 0.53 |
| Clinical Non-Trainee vs Clinical Trainee | -0.756 | **0.01** |
| Staff vs Clinical Trainee | -0.887 | **0.01** |
| Quality of Life |  |  |
| Staff vs Clinical Non-Trainee | -0.344 | 0.09 |
| Clinical Non-Trainee vs Clinical Trainee | -0.189 | 0.45 |
| Staff vs Clinical Trainee | -0.533 | 0.07 |

\*Chi-square test used for statistical comparison

**Supplementary Table 3.** Univariate analysis of factors associated with longitudinal perceived stress.

|  | **Effect Estimate** | **P value** |
| --- | --- | --- |
| Baseline Resilience | -0.84 | <0.001 |
| Baseline Optimism | -0.34 | <0.001 |
| Baseline Emotional Support | -0.62 | <0.001 |
| Baseline Quality of Life | -0.71 | <0.001 |
| Longitudinal Quality of Life | -0.80 | <0.001 |
| Baseline Anxiety/Depression | 1.27 | <0.001 |
| Baseline Body Mass Index | 0.07 | 0.001 |
| Male Gender | -0.94 | 0.002 |
| Mean New York City Case Count Per Period | 0.82 | 0.004 |
| No Positive Corona Virus Nasal PCR at Baseline | 0.91 | 0.114 |
| 2 Week Period Post Positive Corona Virus Nasal PCR | -0.18 | 0.615 |
| 4 Week Period Post Positive Corona Virus Nasal PCR | -0.82 | 0.014 |
| 2 Week Period Post Positive Corona Virus Antibody | -0.36 | 0.191 |
| 4 Week Period Post Positive Corona Virus Antibody | -0.10 | 0.71 |
| Any Period Post Positive Corona Virus Nasal PCR | -0.85 | 0.04 |
| 2 Week Period Post Positive Corona Virus PCR or Antibody | -0.25 | 0.28 |
| Any Period Post Positive Corona Virus Antibody | -0.26 | 0.36 |
| Weight | 0.02 | 0.04 |
| Age | -0.03 | 0.047 |
| Baseline Asthma | 0.89 | 0.045 |
| Baseline Heart Disease | 3.37 | 0.186 |
| Baseline Hypertension | -0.34 | 0.58 |
| Baseline Diabetes | -0.20 | 0.85 |
| Mean Symptomatic Days Per Period | 0.32 | 0.07 |
| Staff vs Clinical Non-Trainee | -0.03 | 0.81 |
| Staff vs Clinical Trainee | 0.43 | 0.15 |
| Clinical vs Clinical Trainee | 0.60 | 0.17 |
| Any Period Post Positive Corona Virus PCR or Antibody | -0.47 | 0.07 |
| Height | -0.03 | 0.08 |
| Mean Days Travelled Per period | -0.09 | 0.58 |
| Days Left Home Per Period | -0.05 | 0.08 |
| No Immune Suppressing Medication at Baseline | -1.80 | 0.17 |
| No Child Care Needs at Baseline | -0.39 | 0.23 |
| Smoking at Baseline | 0.53 | 0.20 |
| Mean Days Hospitalized Per Period | -1.66 | 0.27 |
| Mean Days Treating COVID-19 Positive Patients Per Period | 0.22 | 0.28 |
| Number of Days Left Home | -0.02 | 0.30 |
| Asian vs Black | 0.02 | 0.97 |
| Asian vs Other | -0.34 | 0.47 |
| Black vs Other | -0.36 | 0.55 |
| White vs Asian | 0.01 | 0.99 |
| White vs Black | 0.03 | 0.96 |
| White vs Other | -0.34 | 0.45 |
| No Positive COVID-19 Antibody at Baseline | 0.03 | 0.69 |
| Days Caring for Patients With COVID-19 | 0.01 | 0.73 |
| Interacted With 1-3 People Outside The Home Per Day | 0.01 | 0.97 |
| Interacted With 4-9 People Outside The Home Per Day | 0.01 | 0.96 |
| Interacted With $\geq 10$ People Outside The Home Per Day | -0.11 | 0.64 |
| Mean Days Quarantined Per Period | -0.20 | 0.80 |
| Total Symptomatic Days Per Period | 0.01 | 0.85 |
| Days Working Weighted Based on Patient Exposure | 0.03 | 0.69 |
| Days Hospitalized Per Period | 0.07 | 0.88 |
| Days Quarantined Per Period | -0.03 | 0.90 |
| Mean Number of Days Left the House Per Period | 0.03 | 0.90 |
| Mean Days Working this Period | 0.02 | 0.88 |
| Sum of the Severity of COVID-19 Symptoms Per Period | -0.001 | 0.96 |
| Mean Severity of COVID-19 Symptoms This Period | 0.08 | 0.14 |

**Supplementary Table 4.** Mean HRV parameters stratified based upon emotional support and resilience tertials.

| Parameter | Emotional Support Tertial | Mean SDNN Emotional Support Tertial ms (95% CI) | Resilience Tertial | Mean SDNN Resilience Tertial ms (95% CI) |
| --- | --- | --- | --- | --- |
| MESOR |  |  |  |  |
|  | Low | 42.80 (37.93-47.42) | Low | 45.03 (41.01-48.08) |
|  | Medium | 44.31 (41.32-47.32) | Medium | 42.38 (39.21-45.49) |
|  | High | 43.8 (41.69-46.05) | High | 43.29 (39.83- 46.64) |
| Amplitude |  |  |  |  |
|  | Low | 6.77 (5.66-7.89) | Low | 7.09 (6.44- 7.80) |
|  | Medium | 6.48 (5.79-7.15) | Medium | 5.49 (4.79- 6.19) |
|  | High | 4.85 (4.35-5.38) | High | 4.85 (3.93-5.67) |
| Acrophase |  |  |  |  |
|  | Low | -2.32 (-2.45- -2.20) | Low | -2.38 (-2.46- -2.31) |
|  | Medium | -2.35 (-2.43- -2.27) | Medium | -2.29 (-2.37- -2.20) |
|  | High | -2.52 (-2.61- -2.43) | High | 2.57 (2.75- -2.40) |

**Supplementary Table 5.** Comparison of mean HRV parameters stratified based upon emotional support and resilience tertials.

| Parameter | Emotional Support Tertial Comparisons | *P*-value | Resilience Tertial Comparisons | *P*-value |
| --- | --- | --- | --- | --- |
| MESOR |  |  |  |  |
|  | Low vs Medium | 0.60 | Low vs Medium | 0.10 |
|  | Low vs High | 0.67 | Low vs High | 0.46 |
|  | Medium vs High | 0.78 | Medium vs High | 0.71 |
| Amplitude |  |  |  |  |
|  | Low vs Medium | 0.68 | Low vs Medium | <0.001 |
|  | Low vs High | 0.01 | Low vs High | <0.001 |
|  | Medium vs High | <0.001 | Medium vs High | 0.24 |
| Acrophase |  |  |  |  |
|  | Low vs Medium | 0.70 | Low vs Medium | 0.09 |
|  | Low vs High | 0.004 | Low vs High | 0.048 |
|  | Medium vs High | <0.001 | Medium vs High | <0.001 |

## References

1. Preti E, Di Mattei V, Perego G, et al. The Psychological Impact of Epidemic and Pandemic Outbreaks on Healthcare Workers: Rapid Review of the Evidence. Curr Psychiatry Rep. 07 2020;22(8):43. doi:10.1007/s11920-020-01166-z

2. Bai Y, Lin CC, Lin CY, Chen JY, Chue CM, Chou P. Survey of stress reactions among health care workers involved with the SARS outbreak. Psychiatr Serv. Sep 2004;55(9):1055–7. doi:10.1176/appi.ps.55.9.1055

3. Charney AW, Katz C, Southwick SM, Charney DS. A Call to Protect the Health Care Workers Fighting COVID-19 in the United States. Am J Psychiatry. 10 2020;177(10):900–901. doi:10.1176/appi.ajp.2020.20040535

4. Lai J, Ma S, Wang Y, et al. Factors Associated With Mental Health Outcomes Among Health Care Workers Exposed to Coronavirus Disease 2019. JAMA Netw Open. 03 2020;3(3):e203976. doi:10.1001/jamanetworkopen.2020.3976

5. Liang Y, Wu K, Zhou Y, Huang X, Liu Z. Mental Health in Frontline Medical Workers during the 2019 Novel Coronavirus Disease Epidemic in China: A Comparison with the General Population. Int J Environ Res Public Health. 09 2020;17(18) doi:10.3390/ijerph17186550

6. Rossi R, Socci V, Pacitti F, et al. Mental Health Outcomes Among Frontline and Second-Line Health Care Workers During the Coronavirus Disease 2019 (COVID-19) Pandemic in Italy. JAMA Netw Open. 05 2020;3(5):e2010185. doi:10.1001/jamanetworkopen.2020.10185

7. Zerbini G, Ebigbo A, Reicherts P, Kunz M, Messman H. Psychosocial burden of healthcare professionals in times of COVID-19 - a survey conducted at the University Hospital Augsburg. Ger Med Sci. 2020;18:Doc05. doi:10.3205/000281

8. Morgantini LA, Naha U, Wang H, et al. Factors contributing to healthcare professional burnout during the COVID-19 pandemic: A rapid turnaround global survey. PLoS One. 2020;15(9):e0238217. doi:10.1371/journal.pone.0238217

9. Shaffer F, Ginsberg JP. An Overview of Heart Rate Variability Metrics and Norms. Front Public Health. 2017;5:258. doi:10.3389/fpubh.2017.00258

10. Cohen S, Kamarck T, Mermelstein R. A global measure of perceived stress. J Health Soc Behav. Dec 1983;24(4):385–96.

11. Connor KM, Davidson JR. Development of a new resilience scale: the Connor-Davidson Resilience Scale (CD-RISC). Depress Anxiety. 2003;18(2):76–82. doi:10.1002/da.10113

12. Hahn EA, Devellis RF, Bode RK, et al. Measuring social health in the patient-reported outcomes measurement information system (PROMIS): item bank development and testing. Qual Life Res. Sep 2010;19(7):1035–44. doi:10.1007/s11136-010-9654-0

13. Hays RD, Schalet BD, Spritzer KL, Cella D. Two-item PROMIS® global physical and mental health scales. J Patient Rep Outcomes. 2017;1(1):2. doi:10.1186/s41687-017-0003-8

14. Scheier MF, Carver CS, Bridges MW. Distinguishing optimism from neuroticism (and trait anxiety, self-mastery, and self-esteem): a reevaluation of the Life Orientation Test. J Pers Soc Psychol. Dec 1994;67(6):1063–78. doi:10.1037//0022-3514.67.6.1063

15. Monitor your heart rate with Apple Watch. Accessed October 22nd, 2020, https://support.apple.com/en-us/HT204666

16. New York City Case Count. https://raw.githubusercontent.com/nychealth/coronavirus-data/master/

17. Hirten RP, Danieletto M, Tomalin L, et al. Longitudinal Physiological Data from a Wearable Device Identifies SARS-CoV-2 Infection and Symptoms and Predicts COVID-19 Diagnosis. medRxiv. 2020;doi:https://doi.org/10.1101/2020.11.06.20226803

18. Rutter M. Resilience as a dynamic concept. Dev Psychopathol. May 2012;24(2):335–44. doi:10.1017/S0954579412000028

19. Rutter M. Annual Research Review: Resilience--clinical implications. J Child Psychol Psychiatry. Apr 2013;54(4):474–87. doi:10.1111/j.1469-7610.2012.02615.x

20. Rutter M. Implications of resilience concepts for scientific understanding. Ann N Y Acad Sci. Dec 2006;1094:1–12. doi:10.1196/annals.1376.002

21. Herrman H, Stewart DE, Diaz-Granados N, Berger EL, Jackson B, Yuen T. What is resilience? Can J Psychiatry. May 2011;56(5):258–65. doi:10.1177/070674371105600504

22. Holz NE, Tost H, Meyer-Lindenberg A. Resilience and the brain: a key role for regulatory circuits linked to social stress and support. Mol Psychiatry. 02 2020;25(2):379–396. doi:10.1038/s41380-019-0551-9

23. Kim HG, Cheon EJ, Bai DS, Lee YH, Koo BH. Stress and Heart Rate Variability: A Meta-Analysis and Review of the Literature. Psychiatry Investig. Mar 2018;15(3):235–245. doi:10.30773/pi.2017.08.17

24. Fox S, Lydon S, Byrne D, Madden C, Connolly F, O’Connor P. A systematic review of interventions to foster physician resilience. Postgrad Med J. Mar 2018;94(1109):162–170. doi:10.1136/postgradmedj-2017-135212

25. Mealer M, Conrad D, Evans J, et al. Feasibility and acceptability of a resilience training program for intensive care unit nurses. Am J Crit Care. Nov 2014;23(6):e97–105. doi:10.4037/ajcc2014747

26. Lemaire JB, Wallace JE, Lewin AM, de Grood J, Schaefer JP. The effect of a biofeedback-based stress management tool on physician stress: a randomized controlled clinical trial. Open Med. 2011;5(4):e154–63.

